# First Detection of WHO-validated artemisinin resistance *Plasmodium falciparum* K13 C469Y mutation in patient from Senegal

**DOI:** 10.64898/2026.09.11.26362519

**Authors:** Amy Gaye, Bassirou Ngom, Djiby Sow, Aita Sene, Mariama Toure, Abdoullaye Tine, Yaye Die Ndiaye, Baba Dieye, Khadim Diongue, Younouss Diedhiou, Mame Fama Ndiaye, Mouhamadou Ndiaye, Amadou Moctar Mbaye, Aliou Thiongane, Naby Kaly Standeur, Lamine Ndiaye, Daba Zoumarou, Jules François Gomis, Mame Cheikh Seck, Ibrahima Mbaye Ndiaye, Ibrahima Diallo, ElHadje Doucoure, Ndeye Coumba Diop, Abdoulaye Kane Dia, Aida Sadikh Badiane, Mamadou Alpha Diallo, Awa Bineta Deme, Daouda Ndiaye

## Abstract

Artemisinin-based Combination Therapies (ACTs) remain the first-line treatment for uncomplicated *Plasmodium falciparum* malaria in Senegal. The recent emergence of World Health Organization (WHO)-validated *Pfkelch13* (*K13*) mutations associated with artemisinin partial resistance in Africa highlights the need for continuous molecular surveillance. We conducted a nationwidegenomic surveillance study of 3,800 *P. falciparum* isolates collected from nine malaria-endemic regions of Senegal during 2023-2024. Mutations in the *K13* gene were detected using targeted amplicon deep sequencing, and mutations were independently confirmed by allele-specific quantitative PCR (qPCR). Among all isolates analyzed, a single sample collected in Kafountine, Ziguinchor Region, harbored the WHO-validated K13 C469Y mutation at a allele frequency of approximately 15%, indicating a minority mutant parasite population. Allele-specific qPCR confirmed the presence of the mutant allele, and epidemiologic investigation indicated the patient had no recent travel history outside the study area, suggesting local acquisition. No additional WHO-validated *K13* resistance mutations were detected, and the patient recovered completely following treatment with artemether-lumefantrine. To our knowledge, this finding represents the first detection of the WHO-validated *K13* C469Y mutation in Senegal. Although identified in a single mixed infection and not associated with clinical evidence of ACT failure, the detection of a validated artemisinin resistance mutation prompts deeper investigation and underscores the importance of integrating molecular surveillance into routine malaria control programs to enable early detection of emerging resistance to support evidence-based public health decision-making.

## Background

Malaria remains one of the world’s deadliest infectious diseases, with an estimated 282 million cases and ∼610,000 deaths globally in 2024 (1). More than 94% of cases occurred in sub-Saharan Africa (1), disproportionately affecting children and pregnant individuals. Despite substantial progress over the past 2 decades, recent reports indicate that declines in malaria morbidity and mortality have plateaued, underscoring the need for ongoing surveillance to track trends and ensure that current antimalarial interventions including treatments remain effective (2). ACTs have remained the first-line treatment for uncomplicated Plasmodium falciparum malaria since their adoption by the World Health Organization (WHO) in the early 2000s and have substantially reduced malaria-related morbidity and mortality (3,4). However, their long-term effectiveness is threatened by the emergence of partial artemisinin resistance (ARTR), characterized by delayed parasite clearance that may facilitate selection of partner drug resistance, ultimately compromising ACT efficacy (1,3,5). A major advance in understanding ARTR came with the identification of mutations in the propeller domain of the P. falciparum kelch13 (Pfkelch13, K13) gene, which were shown to be strongly associated with delayed parasite clearance (6). Since then, multiple validated K13 changes, including F446I, N458Y, C469Y, M476I, Y493H, R539T, I543T, P553L, R561H, P574L, C580Y, and R622I, have been classified by the WHO as validated markers of ARTR that are used for genomic surveillance (1). Although ARTR was first identified in Southeast Asia, the independent emergence of validated K13 mutations in several African countries highlighted the importance of molecular surveillance across Africa (7,8). ARTR K13 mutation distribution varies across Africa, with differential frequencies reported for both validated (R622I, R561H, A675V, P574L, C469Y and C580Y) and candidate (C469F, P441L and G449A) variants. R561H and R622I have become established in several East African countries, while C469Y has now been documented in Uganda, Kenya, Tanzania and Ghana. This indicates an expanding geographical distribution of this WHO-validated marker of artemisinin partial resistance (9–12). Among these African resistance alleles,the emergence of the C469Y mutation is particularly noteworthy. Initially detected at very low frequencies in Uganda, this mutation was subsequently shown to be associated with delayed parasite clearance and reduced artemisinin susceptibility and has since been recognized by the World Health Organization as a validated molecular marker of artemisinin partial resistance (1,8). More importantly, genomic analyses demonstrated that C469Y arose on an indigenous African parasite genetic background, highlighting independent evolutionary origins of resistance within Africa rather than transcontinental dissemination (12). This finding fundamentally changes the objectives of malaria surveillance programmes, shifting the focus from detecting imported resistant parasites to identifying the earliest stages of local evolutionary events.

Senegal has made substantial progress in malaria control and is moving toward malaria elimination, making the preservation of ACT efficacy a national public health priority. Although therapeutic efficacy studies (TES) continue to demonstrate excellent clinical performance of ACTs, low-frequency resistant parasite populations may remain undetected by conventional molecular methods. High-depth targeted amplicon sequencing offers improved sensitivity for detecting minority resistance alleles at an early stage of emergence (13).

In this study, we used targeted deep sequencing to characterize the diversity of *K13* mutations in *P. falciparum* isolates collected from passive case detection at health facilities across Senegal during 2023-2024. We further validated any WHO-recognized resistance mutation by allele-specific quantitative PCR and investigated its epidemiologic context through field investigation. Collectively, these complementary approaches provide evidence for detection of the WHO-validated *K13* C469Y mutation in Senegal before any evidence of widespread clinical artemisinin resistance.

## MATERIAL AND METHODS

### Study design

This study was designed as a health facility based, passive case detection, multisite molecular surveillance study conducted in nine regions of Senegal: Kedougou, Tambacounda, Ziguinchor, Sedhiou Diourbel, Kolda, Kaolack, Matam and Thiès from 2023 to 2024 (Figure 1). A total of 3800 patients with confirmed *P. falciparum* infection were enrolled, with 1800 in 2023 and 2000 patients in 2024 from various malaria surveillance sites. These included sentinel sites, routine clinical sites and community-based case management platforms. Sentinel sites contributed standardised clinical and epidemiological data through the national data reporting system, while community sites provided access to peripheral populations not routinely captured in facility-based surveillance. Together, these sites ensured broad representation of the country’s epidemiological strata, from the southeastern areas with high transmission to the western zones with lower transmission (Figure 1).

**Figure 1:**
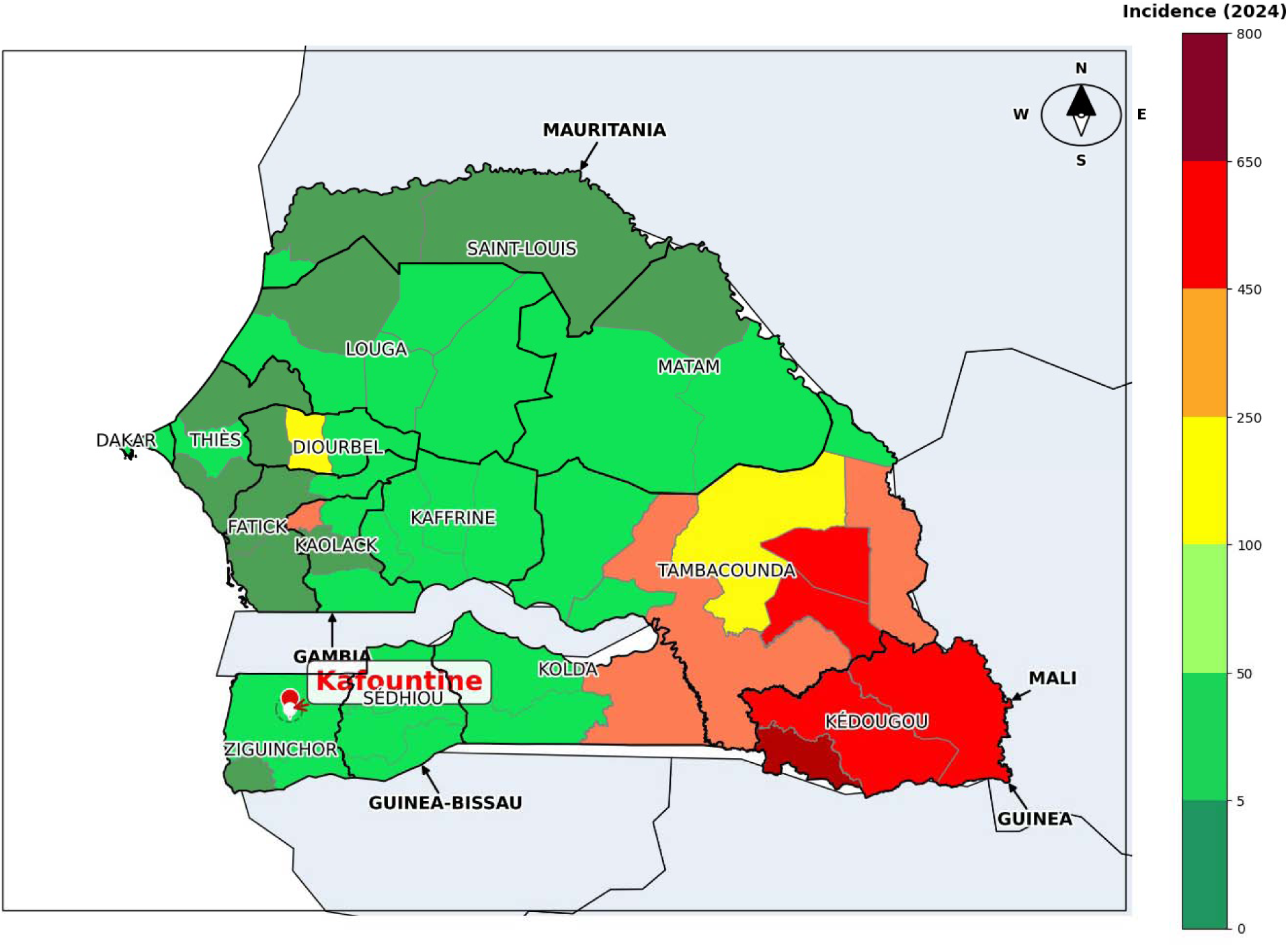
Geographical distribution of the malaria molecular surveillance sites included in this study across Senegal. The map shows the eight regions that were included in the nationwide, cross-sectional, molecular surveillance study that was conducted between 2023 and 2024. Kédougou, Tambacounda, Ziguinchor, Sédhiou, Kolda, Kaolack, Diourbel, Matam and Thiès. Kafountine, in the Ziguinchor Region, is highlighted as it is the location where the WHO-validated K13 C469Y mutation was identified.

### Ethical considerations

This study was approved by the National Ethics Committee of the Ministry of Health in Senegal (Protocol SEN 14/49), the Institutional Review Board of the Harvard T.H. Chan School of Public Health (IRB CR-16330) and conducted in collaboration with the PNLP and regional health authorities. All participants were enrolled after written informed consent, and results were shared with the PNLP to support ongoing surveillance of malaria drug resistance and policy updates.

### Sample collection

Participants were individuals of all ages presenting with fever or history of fever within the previous 48 hours and testing positive for *P. falciparum* by rapid diagnostic test (RDT) or microscopy. Only patients with uncomplicated malaria who provided written informed consent (or assent with parental consent for minors) were included. For each enrolled patient, capillary blood was collected on filter papers (Whatman Protein Saver FTA (Whatman® 3MM CHR CAT N° 3030-662) to obtain dried blood spots (DBS) The samples were stored in plastic bags at room temperature and protected with silica gel desiccant prior to DNA isolation and molecular testing. All samples were coded and anonymous in accordance with the consent process. Standard clinical and demographic information was recorded, including age, sex, date of consultation, previous antimalarial treatment, and geographic coordinates of the health facility or community site.

### Sample Selection for Targeted Deep Sequencing

A total of 3,800 patients with confirmed malaria were included in the study. However it was not possible to sequence all the samples that were collected, and based upon previous empirical data or power calculations using DRpower a subset of representative samples were chosen. This strategy took into account different levels of transmission across each region. This approach kept the study’s geographic and epidemiological diversity while making the most of the available sequencing resources. As a result, 1,828 samples were included in the targeted amplicon deep sequencing analysis.

### Laboratory Procedures

#### DNA extraction

After screening, a total of 1828 samples were subjected to Genomic DNA extracted from dried blood spot (DBS) material using the QIAamp DNA Blood Mini Kit (Qiagen, Valencia, CA, USA).

#### MAD4HatTeR amplicon protocol

Library preparation was conducted following the Paragon Genomics CleanPlex NGS Panel Protocol (version 1.0.1; full protocol and updates available at https://eppicenter.ucsf.edu/resources). This targeted amplicon panel encompasses 276 genomic loci, generating amplicons ranging from 225 to 300 bp. For the present study, we employed the Diversity Module (Primer Pool D1.1), which targets 165 high-diversity loci, as well as the Resistance Module (Primer Pools R1.2 and R2.1), covering 118 loci across 15 antimalarial drug-resistance genes, the *csp* C-terminal region, *hrp2/3* deletion surveillance targets, and non-*falciparum* species detection. Drug-resistance loci included those within *crt*, *mdr1*, *dhfr*, *dhps*, *kelch13*, *coronin*, *exo*, *fd*, *arps10*, and *mdr2* (14).

Two multiplex PCR reactions were performed: the first combining primer pools D1.1 and R1.2, and the second using primer pool R2.1 alone. This dual-reaction design enables full tiling coverage of the genomic regions of interest. Following amplification, PCR products from both reactions were pooled and purified using the bead-based CleanPlex reagent provided by Paragon Genomics. Beads were added at a 1.3X ratio relative to the pooled PCR volume, washed twice with 70% ethanol, and DNA was eluted in 10 μL of TE buffer. Purified amplicons were subsequently subjected to a digestion step to remove non-specific amplification products, followed by an additional bead purification as described. Finally, indexing PCR was performed with 0.5 μM customized unique-dual indexing primers from Paragon Genomics and amplified. Libraries were cleaned to have an amplicon size ∼250 to 350 bp, pooled and sequenced using Illumina MiSeq with 151 bp paired-end.

#### Data Analysis

Bioinformatic processing was performed using the MAD4HatTeR accompanying pipeline for demultiplexing, alignment to the *P. falciparum* 3D7 reference genome, and within-sample allele frequency estimation at each targeted locus. The platform reliably detects minority variants at within-sample allele frequencies as low as 1% in high-density specimens, enabling identification of sub-patent resistance genotypes in polyclonal infections. To ensure robustness of variant calls, raw sequencing reads were independently re-analyzed using the MaRS bioinformatic pipeline (15), which performs independent demultiplexing, alignment, and variant calling. Read alignments spanning the *K13* propeller domain were additionally visualized in Geneious Prime to allow manual inspection for sequencing artifacts, strand bias, and positional errors prior to reporting any variant as a true positive.

#### Allele-specific qPCR assay design

To validate the K13 C469Y variant identified by targeted amplicon deep sequencing independently, a quantitative PCR assay using an allele-specific hydrolysis probe (TaqMan) was designed to distinguish between the wild-type (C469; TGC) and mutant (Y469; TAC) alleles. The assay targeted the nucleotide substitution that causes the amino acid change from C469 to Y469 within the K13 propeller domain.The primer and probe sequences were designed to amplify a 92 bp fragment spanning codon 469. The forward primer sequence was 5′-GGTGGATTTGATGGTGTGAAAT-3′ and the reverse primer sequence was 5′-TTGGTAGACATAGGTGTACCAA-3′; both had a predicted melting temperature of approximately 62°C. Two allele-specific Locked Nucleic Acid (LNA) hydrolysis probes were designed to maximize discrimination between the reference and mutant alleles. The reference-specific probe (C469, G allele) sequence was 5′-ACGCCA+G+C+ATT+GT-3′, and the mutant-specific probe (Y469, A allele) sequence was 5′-ACG+CCA+G+T+AT+TG-3′. The predicted mismatch melting temperatures (53.0°C and 55.9°C, respectively) ensured high allele specificity. Relative quantitative PCR were assessed using a real-time PCR machine (ABI 7500). Briefly, qPCR was carried out in 15μl volumes in a 96-well plate containing 10 μl Taqman environmental master mix, 0.5 μl of each forward and reverse primer, and probe, 3 μl H O, and 5 μl template DNA. Amplifications were performed under the following conditions: 98 °C for 3 min, followed by 40 cycles of 95 °C for 10 s and 64 °C for 20 s. Amplifications were run in triplicate.

#### Field Epidemiological Investigation

Following identification of C469Y in a sample from the Kafountine health post (Diouloulou District, Ziguinchor Region), a two-person investigation team comprising a field technician and a research biologist traveled to the site to interview the index patient, document the clinical history, and collect a detailed travel history to assess whether the infection was locally acquired or potentially imported (Figure 2).

**Figure 2.**
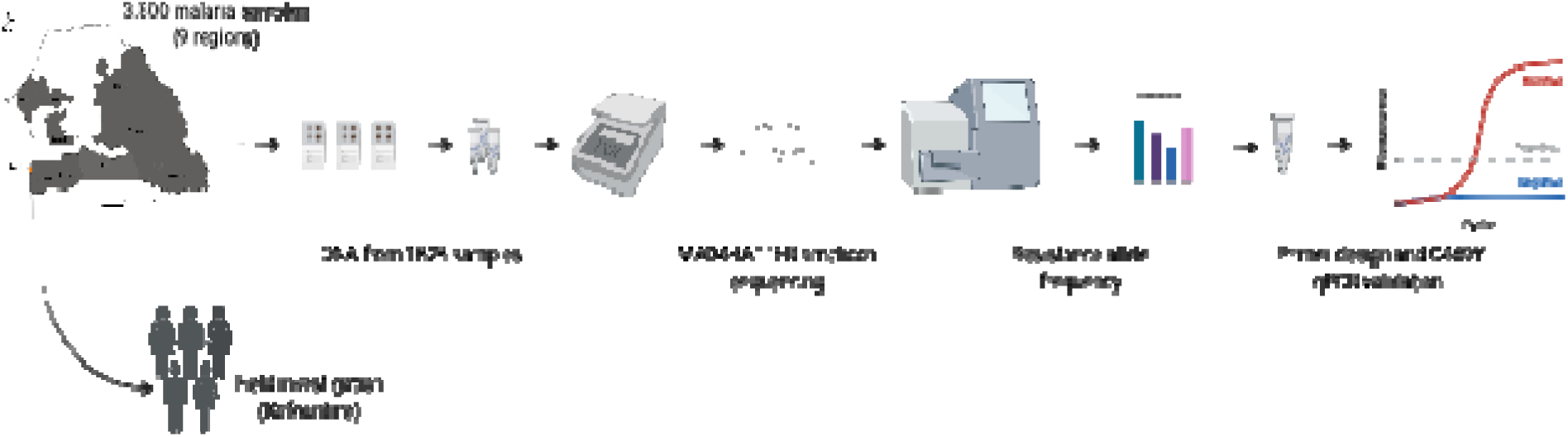
Study design and analytical workflow for national molecular surveillance of *P. falciparum K13* polymorphisms and independent confirmation of the C469Y mutation in Senegal. Image created with BioRender.com.

Figure 2 summarizes the overall study design and analytical workflow used to investigate *K13* polymorphisms in *P. falciparum* isolates collected through Senegal’s national surveillance program. The workflow integrates field sampling, genomic analysis, bioinformatic processing, and molecular validation into a comprehensive surveillance framework.

## Results

From this national surveillance system, only a single individual harbored a *P. falciparum* infection that showed evidence of a WHO-validated K13 ART-R mutation. A single patient from Kafountine, in southern Senegal, with a mixed genome infection, harbored the WHO-validated C469Y mutation at an estimated read count variant allele frequency of ∼15%. The mutation was detected using the MAD4HatTeR amplicon sequencing strategy (Figure 4) then independently confirmed by allele-specific quantitative PCR (Figure 5). Case investigation and epidemiological investigation revealed that the patient had no recent travel history outside the study area, suggesting that the infection was likely acquired locally.

### 1 Sequencing Quality Metrics

The K13 C469Y mutation was supported by high sequencing depth, 594 of 3,923 reads covering codon 469 (depth = 3,923×), corresponding to a variant allele frequency (VAF) of ∼15 % (Supplementary Table S1). This coverage depht provided strong confidence in the detection of the variant.

Sequencing depth across the entire *K13* amplicon was consistently high, ranging from several thousand to over 80,000 reads depht depending on the genomic position. This ensured robust variant calling throughout the target region (Supplementary Figure S2). The underlying G:A transversion converts codon TGC (cysteine) to TAC (tyrosine) at propeller-domain position 469 of *K13*. Independent bioinformatic re-analysis of the raw sequencing reads using the MaRS pipeline yielded concordant results, confirming that the detection was not an artifact specific to the primary analysis pipeline (Supplementary Table S2) . Visual inspection of read alignments in Genius Prime corroborated the finding, revealing the G to A transversion at nucleotide position 1406 across multiple independent high-quality reads, with no evidence of strand bias, homopolymer-associated error, or systematic misalignment at the locus.

### 2 National distribution of *K13* polymorphisms detected in isolates collected in Senegal between 2023 and 2024

At the national level, the *K13* locus remained wild-type throughout the study period. Nearly all of the investigated codons were represented exclusively by the reference (wild-type) allele in both 2023 and 2024 (Figure 3). In 2023, only one mixed genotype was identified at the A578S position, accounting for 0.13% of cases (1/795). K13 A578S is a common mutation found in Asia and Africa that is not associated with ART-R. In 2024, a single mixed genotype carrying the WHO-validated C469Y mutation was identified at a frequency of 0.05% (1/1,828).

**Figure 3.**
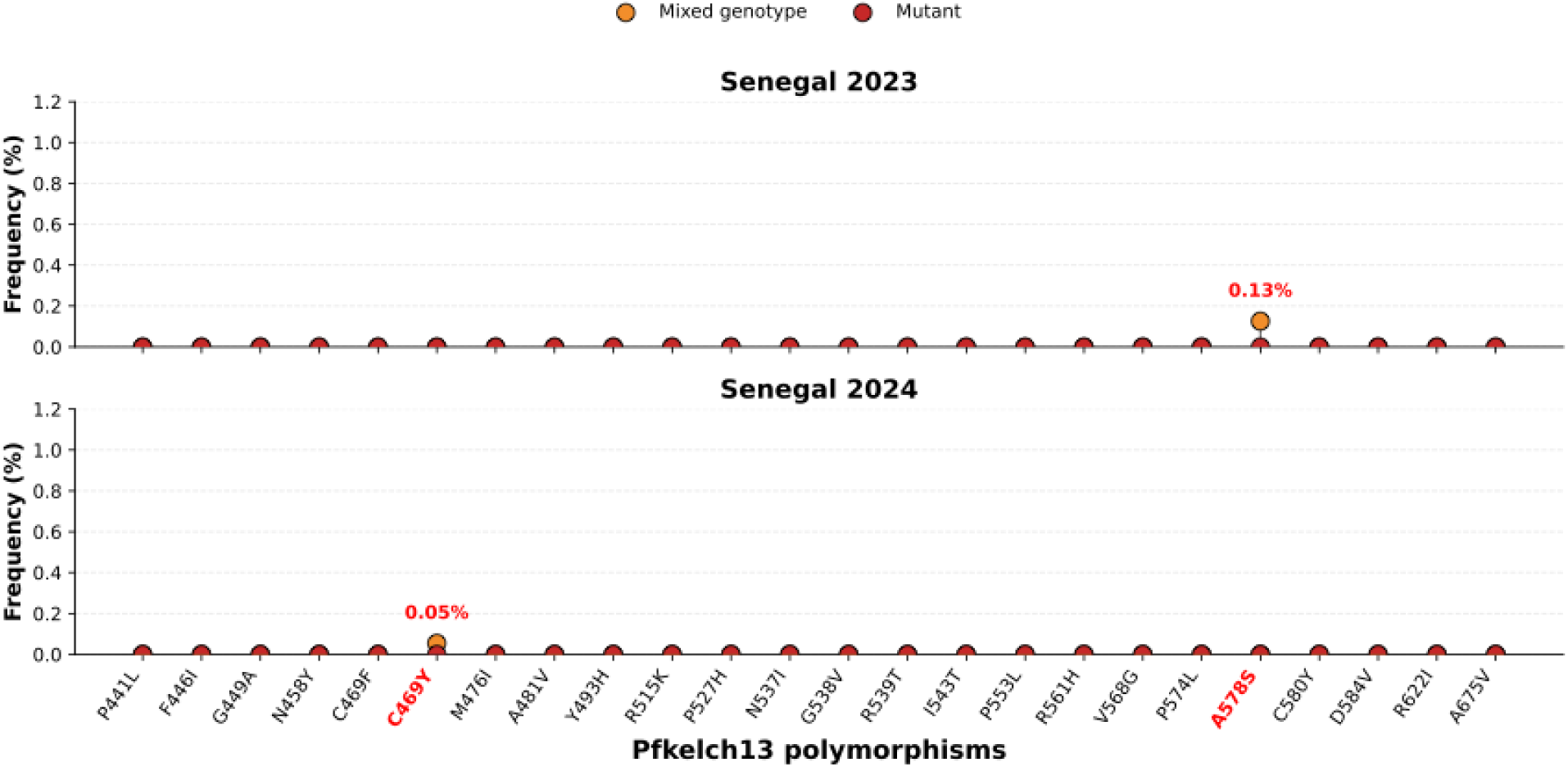
Allele frequencies of K13 polymorphisms among *P. falciparum* isolates from Senegal, 2023 and 2024. This Figure shows the national distribution of *K13* polymorphisms detected in *P. falciparum* isolates collected in Senegal during 2023 (upper panel) and 2024 (lower panel). In 2023, a single mixed genotype was identified at codon A578S, corresponding to 0.13% (1/794) of successfully genotyped isolates. In 2024, a single isolate carrying the WHO-validated C469Y mutation was detected at a frequency of 0.05% (1/1828).

Overall, these findings indicate an overwhelming predominance of wild-type *K13* alleles across Senegal and provide no evidence of widespread artemisinin resistance. However, the detection of the C469Y mutation is important from an epidemiological perspective because the WHO recognizes it as a validated molecular marker of partial ART-R, thus warrants increases surveillance and vigilance. Although it was identified in only one infection and was present as a mixed genotype, this finding suggests the presence of a minority parasite subpopulation that is resistant, rather than an established resistant lineage.

### 3 Allelic frequency of the K13 C469Y mutation based on read counts in a *Plasmodium falciparum* isolate

Allele frequency analysis demonstrates that the C469Y mutation was present as a minor parasite subpopulation, representing approximately 15% of sequencing reads, whereas the wild-type allele accounted for the remaining 85%. All other investigated K13 polymorphisms were exclusively represented by the reference allele, indicating the absence of additional nonsynonymous variants within this isolate (Figure 4).

**Figure 4.**
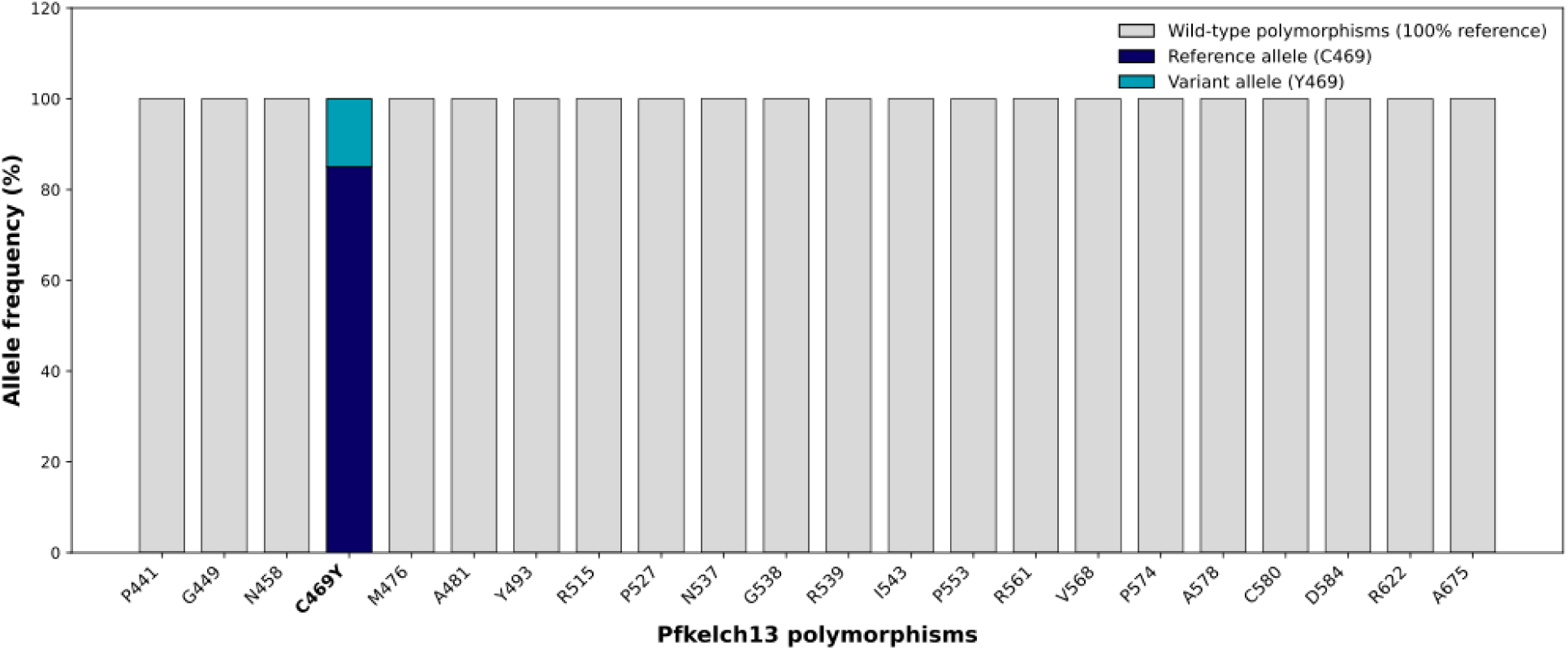
Allele frequency profile of K13 polymorphisms for the *P. falciparum* sample that carried a C469Y mutation. This figure shows the frequencies of the different alleles of all the K13 polymorphisms investigated in the isolate which the C469Y mutation was detected during the national survey. Each bar represents the allele composition at a given K13 codon. Grey bars indicate polymorphic positions entirely represented by the wild-type allele (100% reference). The C469Y position is displayed separately and shows the coexistence of 85% reference (C469) and 15% mutant (Y469) alleles. This is consistent with a mixed infection within the same sample.

### 4 Additional drug-resistance haplotype profile in the C469Y-Positive isolate

To further characterize the genetic background of the isolate carrying the K13 C469Y mutation, we examined haplotypes at additional antimalarial drug-resistance loci targeted by the amplicon sequencing panel (Supplementary Table S1).The isolate carried the Pfcrt CVIET haplotype, defined by the amino acid residues at positions 72–76, at a frequency of 96.76%, whereas CVMNK was detected at 3.24%. At Pfdhfr, the IRN haplotype, corresponding to the N51I, C59R, and S108N combination, was detected at a frequency of 100%, while ICN was absent (0%). For Pfmdr1, the NYD haplotype (N86-Y184-D1246) was present at a frequency of 100%, whereas NFD was not detected (0%). At Pfdhps, the key resistance-associated positions 540 and 580 were represented exclusively by the wild-type alleles (100%).

### 5 independent confirmation by allele-specific qPCR

The allele-specific qPCR assay independently confirmed the presence of the *Pfkelch13* C469Y mutation initially detected by targeted deep amplicon sequencing. As shown in Figure 5, the reference allele was amplified at Ct = 28.1, while the mutant allele was amplified at Ct = 32.9. The wild-type allele amplified approximately 4.8 PCR cycles earlier than the mutant allele, indicating that the wild-type parasite population was substantially more abundant within the infection. Because each PCR cycle represents an exponential increase in DNA amplification, this Ct difference is consistent with the mutant allele being present as a minor parasite subpopulation rather than as the dominant genotype.These findings closely agree with the deep sequencing results, which estimated a variant allele frequency (VAF) of approximately 15% for the C469Y mutation.

**Figure 5.**
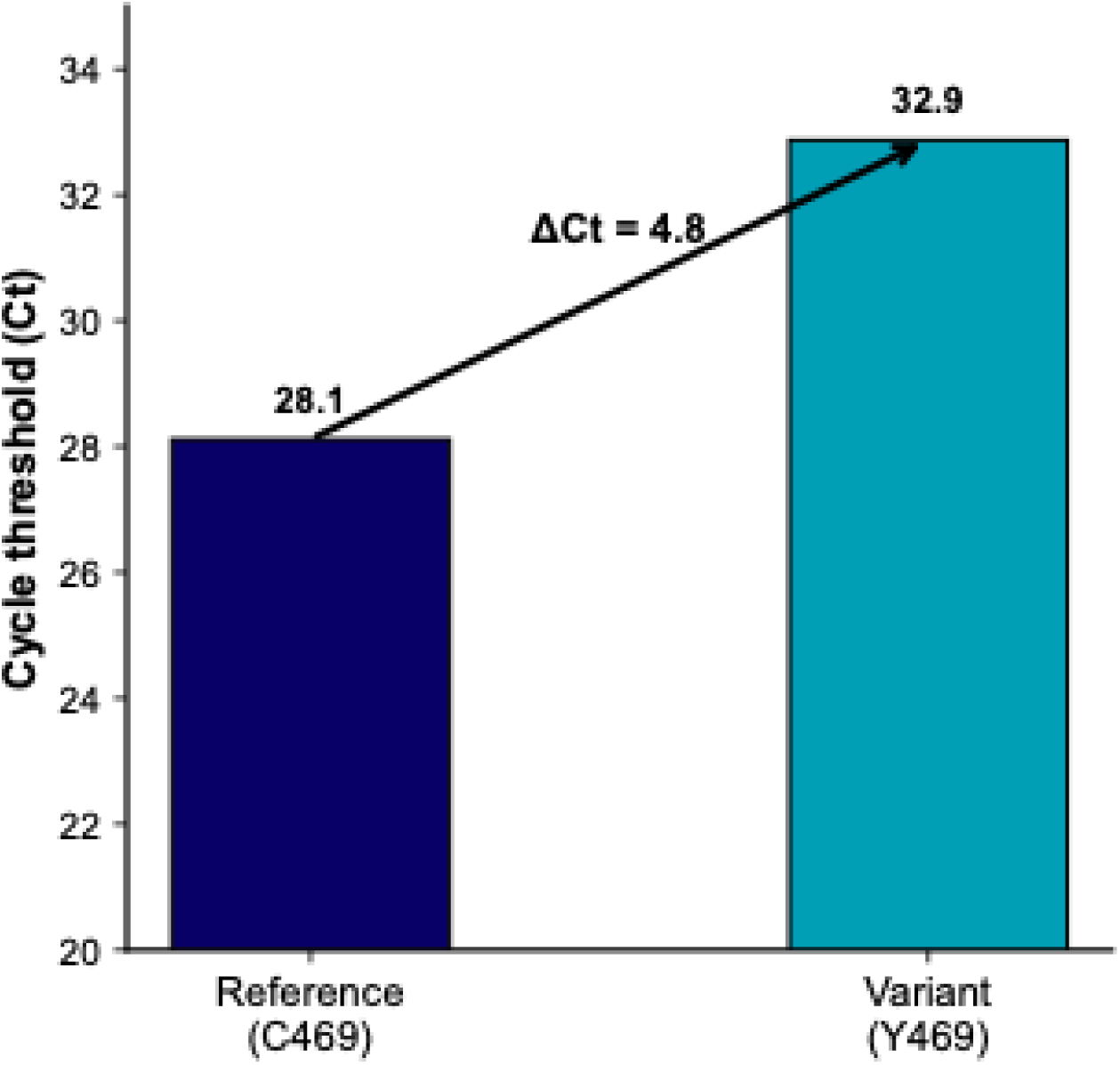
Independent confirmation of the K13 C469Y mutation by allele-specific TaqMan quantitative PCR. The wild-type allele (C469) amplified at a mean cycle threshold (Ct) of approximately 28.1, whereas the mutant allele (Y469) amplified at a mean Ct of approximately 32.9, corresponding to a ΔCt of 4.8 cycles.

### 6 Field Epidemiological Investigation

The patient had a fever of 104°F, headache, and body aches with uncomplicated malaria. Clinical follow-up diagnosed *P. falciparum* infection was confirmed by two different tests: a malaria rapid diagnostic test (RDT) and a test that uses light to look for parasites. During the medical interview, the patient said they hadn’t traveled outside of the area or the surrounding area in the past few years, consistent with a locally obtained infection. The patient received artemether-lumefantrine (Coartem®) according to the Senegalese National Malaria Control Programme (NMCP) treatment guidelines and recovered completely without any problems.

### Disucussion

This study reports, to our knowledge, the first detection of the WHO-validated *Pfkelch13 C469Y* mutation associated with artemisinin partial resistance in Senegal.Using nationwide targeted deep amplicon sequencing, we identified a single isolate carrying the C469Y mutation among 1,828 successfully sequenced *P. falciparum* isolates collected during 2023 and 2024. At the population level, this corresponded to an extremely low prevalence (0.05% among isolates analyzed in 2024), whereas within the infected individual the mutant allele represented approximately 15% of the parasite population, indicating a minority resistant clone. The finding was independently confirmed by allele-specific TaqMan quantitative PCR and supported by field epidemiological investigation indicating probable local acquisition. Although identified in only one patient, this finding represents an important molecular milestone because it extends the geographical distribution of a WHO-validated artemisinin resistance mutation in Senegal and highlights the value of genomic surveillance for detecting the earliest signals of resistance emergence.The emergence of validated *Pfkelch13* mutations has fundamentally changed the epidemiology of artemisinin resistance in Africa. Initially, resistance was considered to be confined to the Greater Mekong Subregion following the discovery of *PfKelch13* mutations associated with delayed parasite clearance (6). However, studies performed over the last few years have demonstrated that Africa has become an independent centre for the evolution of artemisinin resistance. The first evidence came from Rwanda, where the R561H mutation emerged independently on an African parasite genetic background and was associated with delayed parasite clearance following ACT treatment (7). Similar observations were subsequently reported in Uganda, where validated mutations including A675V and C469Y were associated with reduced artemisinin susceptibility and delayed parasite clearance (8). More recent genomic studies further demonstrated that these mutations evolved independently within African parasite populations rather than being introduced from Southeast Asia (12). Our findings therefore support the growing evidence that validated *Pfkelch13* resistance mutations can arise independently across geographically distinct regions of Africa, emphasizing the importance of continued molecular surveillance throughout the continent.

A particularly important aspect of our study is that C469Y was detected as a minority parasite population, representing approximately 15% of the parasites within the infection. The remaining parasite population retained the wild-type allele, indicating that the resistant clone had not yet become dominant. This observation is consistent with current models of resistance evolution, in which drug-resistant parasites initially emerge as minority variants within mixed infections before expanding under sustained antimalarial drug pressure (2,16,17). Similar evolutionary trajectories have previously been described for resistance to chloroquine and sulfadoxine-pyrimethamine, where resistant alleles remained rare before progressively increasing in frequency and ultimately becoming fixed in parasite populations (18–20). Our findings suggest that the C469Y mutation detected in Senegal may represent one of the earliest detectable stages of artemisinin resistance evolution.

Importantly, the detection of C469Y should not be interpreted as evidence that artemisinin resistance is currently established in Senegal. Only one isolate carried a validated resistance mutation, no additional WHO-validated *Pfkelch13* mutations were identified, and the infected patient responded successfully to artemether-lumefantrine with complete clinical recovery. Furthermore, therapeutic efficacy studies conducted by the Senegalese National Malaria Control Programme continue to demonstrate excellent efficacy of ACTs. Together, these observations indicate that ACTs remain highly effective in Senegal and that there is currently no evidence supporting widespread clinical artemisinin resistance. Nevertheless, experiences from Southeast Asia and East Africa have shown that validated resistance mutations may circulate at very low frequencies for several years before becoming sufficiently prevalent to affect parasite clearance or therapeutic efficacy (5–7). Consequently, the detection of C469Y should be regarded as an early warning signal rather than an indication of imminent treatment failure.

Thus, the additional resistance-associated haplotypes observed in this isolate provide further genetic context for the C469Y-containing parasite and suggest a genetic background shaped by previous antimalarial drug selection

The epidemiological investigation provides additional support for the public health significance of this observation. The absence of recent travel history strongly suggests that the infection was locally acquired, although this alone cannot demonstrate that the mutation emerged de novo in Senegal.Whole-genome sequencing and population genomic analyses will be required to determine whether the detected parasite represents an independent evolutionary event or belongs to a lineage circulating more broadly in West Africa. Nevertheless, the integration of molecular findings with epidemiological investigation considerably strengthens the interpretation of this study and illustrates the importance of combining laboratory surveillance with field investigations when validated resistance mutations are detected.

Our study has several strengths. It was conducted within a nationwide molecular surveillance programme covering multiple epidemiological settings across Senegal and combined high-resolution molecular detection with independent qPCR confirmation and epidemiological investigation. This integrated approach substantially increases confidence in the reported finding and provides one of the earliest documented detections of a WHO-validated artemisinin resistance mutation in West Africa. However, several limitations should also be acknowledged. The mutation was detected in only a single isolate, precluding estimation of its prevalence or transmission dynamics. In addition, phenotypic assays such as the ring-stage survival assay were not performed, and whole-genome sequencing was unavailable to investigate the genetic background of the resistant parasite. Continued longitudinal surveillance will therefore be essential to determine whether C469Y remains an isolated observation or increases in frequency over time.

## Conclusion

This study reports the first detection of the WHO-validated *Plasmodium falciparum kelch13* C469Y mutation associated with artemisinin partial resistance in Senegal. Using high-resolution targeted amplicon deep sequencing, we identified the mutation as a minority parasite clone with a allele frequency of approximately 15%, a finding that was independently confirmed by allele-specific qPCR and supported by epidemiological investigation indicating probable local acquisition. Although identified in only a single infection and in the absence of evidence for widespread artemisinin resistance or reduced ACT efficacy, the detection of a validated resistance allele represents an important early molecular warning signal.These findings highlight the value of integrating high-resolution genomic surveillance into routine malaria control programmes to detect emerging resistance before measurable clinical or epidemiological consequences become apparent. The combination of deep sequencing, molecular validation, and field epidemiology provides a robust framework for identifying rare resistance variants at the earliest stages of their evolution. As artemisinin resistance continues to emerge independently across Africa, proactive genomic surveillance will become increasingly important for safeguarding the long-term efficacy of ACTs and informing evidence-based malaria control and elimination strategies. Continued longitudinal surveillance and population genomic investigations will now be essential to determine whether the C469Y mutation remains an isolated event or represents the beginning of an emerging resistant lineage in Senegal and more broadly in West Africa.

## SUPPORTING INFORMATION

**S1 Figure. Comprehensive within-host resistance genotype profile of the Plasmodium falciparum isolate harboring the *Pfkelch13 C469Y* mutation .** Each horizontal bar corresponds to a drug resistance-associated polymorphism included in the targeted amplicon sequencing panel, covering the *Pfkelch13*, *Pfcrt*, *Pfmdr1*, *Pfdhfr*, and *Pfdhps* genes. Variants are classified according to their within-host allele frequency as wild-type (blue), major mutant allele (green; allele frequency ≥95%), or minor mutant allele (pink; allele frequency <95%).

**S2 Figure. Sequencing depth across the sample with Pfkelch13 C469Y mutation generated using the MAD4HatTeR targeted deep sequencing.** This figure shows the sequencing depth obtained across the Pfkelch13 amplicon using the MAD4HatTeR targeted amplicon deep sequencing assay. The x-axis shows the position of the genomic region that has been amplified, and the y-axis shows the number of times that the region has been read. The number of reads is different in each area of the amplicon. In areas with few reads, there are a few thousand reads. In areas with many reads, there are more than 80,000 reads dephts. The highest number of sequencing reads is found in the middle of the amplicon, while the ends have fewer reads.

**S1 Table.** Complete list of the target gene with the allele profile and read count from sample that carried a C469Y mutation using MADHATTER pipeline

**S2 Table.** list of the variant allele frequencies and all the genomic information about the sample with the C469Y mutation using MaRS pipeline.

## Data Availability

All data produced in the present work are contained in the manuscript

## ACKNOWLEDGMENTS

We thank the health workers, community health agents, and PECADOM and PECADOM-Plus supervisors at all sites for helping with sample collection and participant enrollment. We are thankful to the Programme National de Lutte contre le Paludisme for its help with the surveillance program, and to the Regional Health Directorate of Ziguinchor for making the field investigation possible. We also thank the laboratory team at CIGASS for extracting DNA, preparing libraries, and sequencing.

## AUTHOR CONTRIBUTIONS

### FUNDING

This work was supported by the Gates Foundation through a grant to CIGASS for the project “Integrating Genomic Data into Malaria Surveillance and Decision-Making in Senegal.” The funders had no role in study design, data collection, analysis, interpretation, or the decision to publish.

### DATA AVAILABILITY

The raw targeted amplicon sequencing data generated from the *Plasmodium falciparum* isolate carrying the *Pfkelch13* C469Y mutation have been deposited in the National Center for Biotechnology Information (NCBI) Sequence Read Archive (SRA) under BioProject accession number PRJNA1484980 (Temporary Submission ID: SUB16303876). The dataset is currently under embargo and is scheduled for public release on September 30, 2026, or upon publication of this article, whichever occurs first. Access to the data can be provided to the journal editors and reviewers upon request during the peer-review process.

