## Supplementary Figure for "First Detection of WHO-validated artemisinin resistance *Plasmodium falciparum* K13 C469Y mutation in patient from Senegal": Supplementary Figure.docx

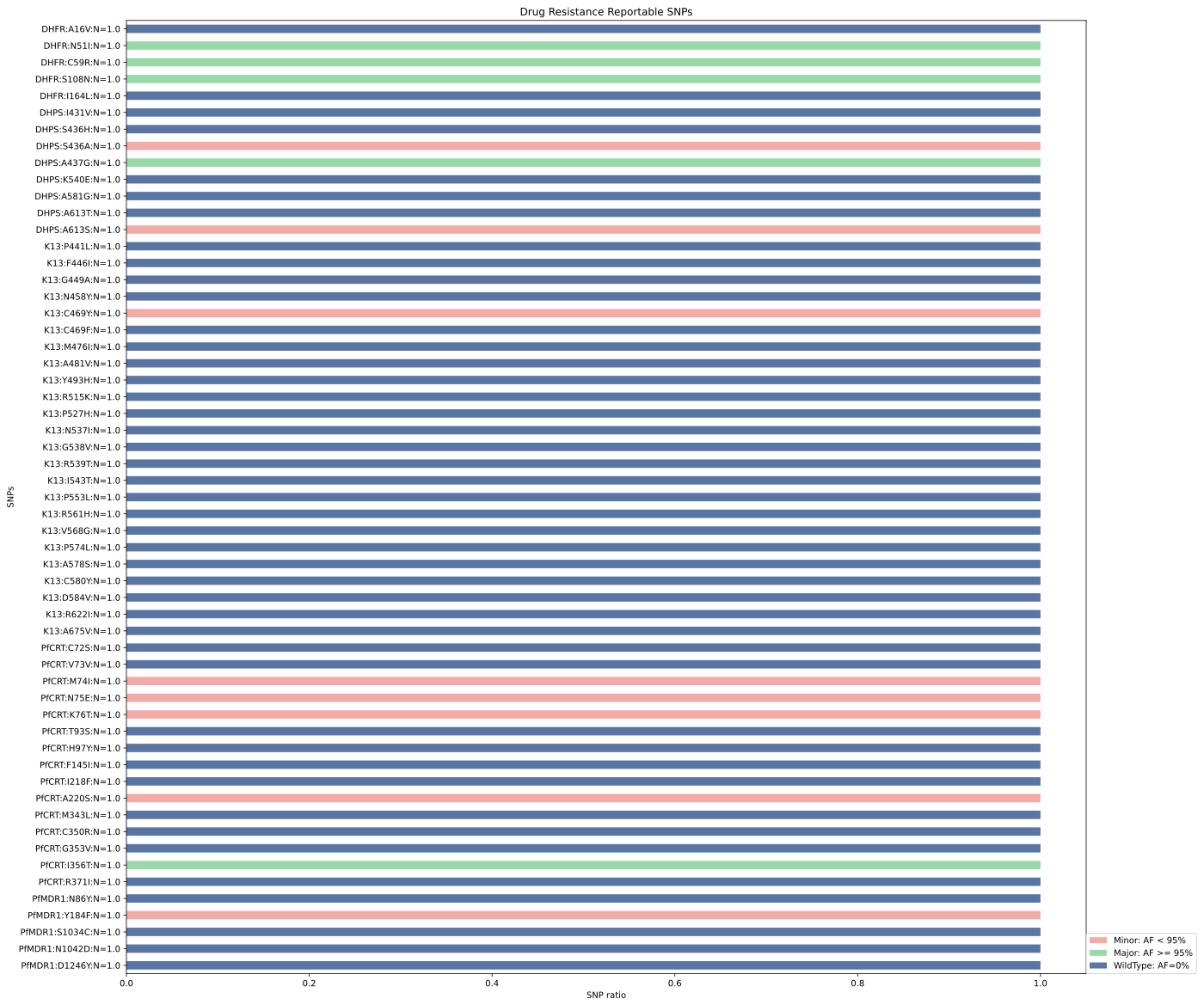

**Appendix Figure S1. Comprehensive within-sample resistance genotype profile of the P. falciparum isolate harboring the Pfkelch13 C469Y mutation**. Each horizontal bar corresponds to a drug resistance-associated polymorphism included in the targeted amplicon sequencing panel, covering *Pfkelch13*, *Pfcrt*, *Pfmdr1*, *Pfdhfr*, and *Pfdhps*. Variants are classified according to within-host allele frequency as wild-type, major mutant allele (allele frequency ≥95%), or minor mutant allele (allele frequency <95%).

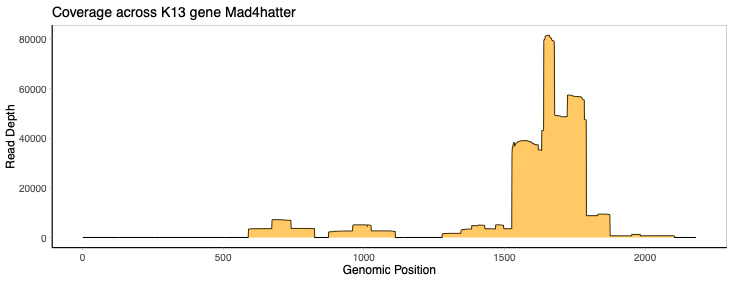

**Appendix Figure S2**. Sequencing depth across the Pfkelch13 amplicon in the sample carrying the C469Y mutation. Sequencing depth varied across the amplicon, ranging from several thousand reads at lower-coverage positions to more than 80,000 reads at higher-coverage positions.
